# Trends in HIV incidence and mortality in Brazil from 2000 to 2022: A Joinpoint Regression Analysis

**DOI:** 10.1101/2025.08.12.25333271

**Authors:** Ketyllem Tayanne da Silva Costa, Yago Tavares Pinheiro, Fábia Barbosa de Andrade, Richardson Augusto Rosendo da Silva

**Affiliations:** Master in Public Health. Department of Public Health, Federal University of Rio Grande do Norte, Rio Grande do Norte, Natal, Brazil; PhD in Public Health. Department of Public Health, Federal University of Rio Grande do Norte, Natal, Brazil; PhD in Health Sciences. Department of Nursing, Federal University of Paraíba, João Pessoa, Paraíba, Brazil; PhD in Health Sciences. Department of Public Health, Federal University of Rio Grande do Norte, Rio Grande do Norte, Natal, Brazil

## Abstract

The present study aims to analyze temporal trends in HIV incidence and mortality in Brazil between 2000 and 2022, using the Joinpoint® software. This ecological study is based on retrospective secondary data collected in October 2024 from the Notifiable Diseases Information System, the Laboratory Test Control System, and the Mortality Information System. Statistical analyses were performed using the Joinpoint Regression Program® (version 4.9.1.0, National Cancer Institute). Ethical approval from a Research Ethics Committee was not required. The North and Northeast regions showed a gradual increase throughout the analyzed time series. The Southeast region, which started in 2000 with the highest incidence rate of 29 cases per 100,000 inhabitants— approximately four times higher than the rates in the North and Northeast regions during the same period—presented one of the lowest rates in the country by 2022. Regarding the HIV mortality rate in Brazil, there was an upward trend in the curve until 2016. On the other hand, the Southeast region was the only one exhibiting line segments with decreasing or stationary Annual Percent Changes throughout the analyzed period, indicating a positive trend in reducing HIV mortality, in contrast to the other Brazilian regions.

## Introduction

The Global Health Financing 2015 report by the Institute for Health Metrics and Evaluation recorded an investment of US$109.8 billion in the fight against HIV/AIDS between 2000 and 2015. However, this investment plateaued after 2010 [1]. Furthermore, it is important to consider that people living with HIV (PLHIV) are particularly vulnerable to opportunistic infections that frequently result in hospitalizations, all of which are fully financed by Brazilian Unified Health System (SUS) [2].

Thus, HIV/AIDS represents a significant public health issue that imposes a high financial burden on the SUS, given that the system invests in prevention, diagnosis, and treatment measures, which can be provided both in specialized clinical centers and through Primary Health Care [2]. In 2019, the Joint United Nations Programme on HIV/AIDS (UNAIDS) estimated 38 million people living with HIV (PLHIV) worldwide, with approximately 920,000 in Brazil alone. That same year, Brazil reported around 48,000 new cases and 14,000 AIDS-related deaths [3].

Due to its epidemiological relevance and the high costs imposed on the health system, the fight against HIV/AIDS was included in the Millennium Development Goals (MDGs), established by United Nations (UN) member countries in 2000, with targets to be achieved by 2015. During this period, Brazil made progress in reducing the HIV incidence. However, the country still faces significant challenges, particularly in terms of access to treatment, falling short of the ideal scenario [4].

As a continuation of these global commitments, the 90-90-90 targets were established for the year 2020, aiming for 90% of people living with HIV (PLHIV) to be diagnosed, 90% of those diagnosed to be on antiretroviral therapy (ART), and 90% of those on ART to achieve viral suppression. Reaching and sustaining these targets is believed to be crucial to ending the HIV/AIDS epidemic globally [5].

To date, ecological studies on HIV infection in Brazil have predominantly focused on specific populations, such as youth, women, or men who have sex with men, or have been limited to specific regions or municipalities [6–9]. In this context, the present study stands out as the first to analyze temporal trends in HIV incidence and mortality considering all reported cases across the entire national territory, without restrictions regarding population profile or geographic location.

This broad approach provides a comprehensive and up-to-date view of the epidemic’s dynamics in Brazil, offering novel insights into the spatiotemporal patterns of the disease at the national level and supplying valuable evidence to inform more equitable and targeted public health policies.

In this context, the present study aims to analyze temporal trends in HIV incidence and mortality in Brazil, between 2000 and 2022, using the Joinpoint® software.

## Methods

### Study Design

This is an ecological study based on retrospective secondary data collected from the Notifiable Diseases Information System (SINAN), the Laboratory Test Control System (SISCEL), and the Mortality Information System (SIM), which are integrated into the platform of the Department of Informatics of the Unified Health System (DATASUS), an agency managed by the Brazilian Ministry of Health.

### Geographic Scope and Population

The study was conducted using data from Brazil, the largest country in South America, with a territorial area of 8,509,379.576 km² [10,11]. The country is divided into five regions—North, Northeast, Southeast, South, and Central-West—comprising a total of 26 states plus the Federal District. The country has an estimated population of 212,583,750, Human Development Index (HDI) of 0.786 and a primary health care coverage rate of 98.16% [12–14]. The population data used refer to the year 2011, as it represents the midpoint of the study period. These demographic characteristics make Brazil a country of great relevance to epidemiology and public health.

### Variables

The data refer to HIV infection and mortality in Brazil during the period from 2000 to 2022, which constitute the dependent variables of the study. The independent variables used were year and region.

### Inclusion and exclusion criteria

All newly reported cases and deaths recorded by health services in the SINAN and SIM systems from the five Brazilian regions between 2000 and 2022 were included. The analysis corresponds to this period due to the beginning of the National STD/AIDS Policy in Brazil until the last year available in the information system. Data with missing values were not considered; in such cases, the mean of the preceding and succeeding values was used.

### Statistical analysis

Data were organized and tabulated by region using the Statistical Package for the Social Sciences (SPSS, version 25, IBM). HIV incidence (1) and mortality (2) rates were calculated based on the number of newly reported cases and recorded deaths, respectively, divided by the resident population for the same period and multiplied by 100,000 inhabitants.

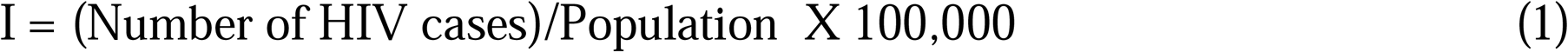

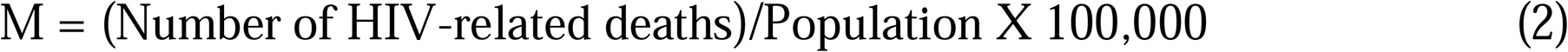

Statistical analysis was performed using the Joinpoint Regression Program® (version 4.9.1.0, National Cancer Institute - NCI). This software conducts regression by identifying inflection points where significant changes in trend occur over the analyzed period, fitting a series of connected linear segments on a logarithmic scale.

Joinpoint® calculates the Annual Percent Change (APC) for each identified segment, as well as the Average Annual Percent Change (AAPC) for the entire period, serving as a summary measure of the trend. Both metrics use 95% confidence intervals (95% CI) and statistical significance tests (p < 0.05). The final model selection was based on the Monte Carlo permutation test.

### Ethical considerations

As this study used aggregated, non-sensitive data without individual information and that are publicly available, ethical approval by the Research Ethics Committee was not required, in accordance with the guidelines established by Resolution No. 510/2016 [15].

## Results

Figure 1a shows the trend analysis of HIV incidence in Brazil conducted using Joinpoint for the period from 2000 to 2020. The results indicate an upward curve until 2012, reaching a peak in 2013 with 23 infected individuals per 100,000 inhabitants. Subsequently, there is a decreasing period, with the lowest rate recorded in 2020—the first year of the COVID-19 pandemic—at 16 diagnoses per 100,000 inhabitants (APC = −6.37). Toward the end of the analyzed period, cases began to rise again (APC = 6.10).

**Fig. 1.**
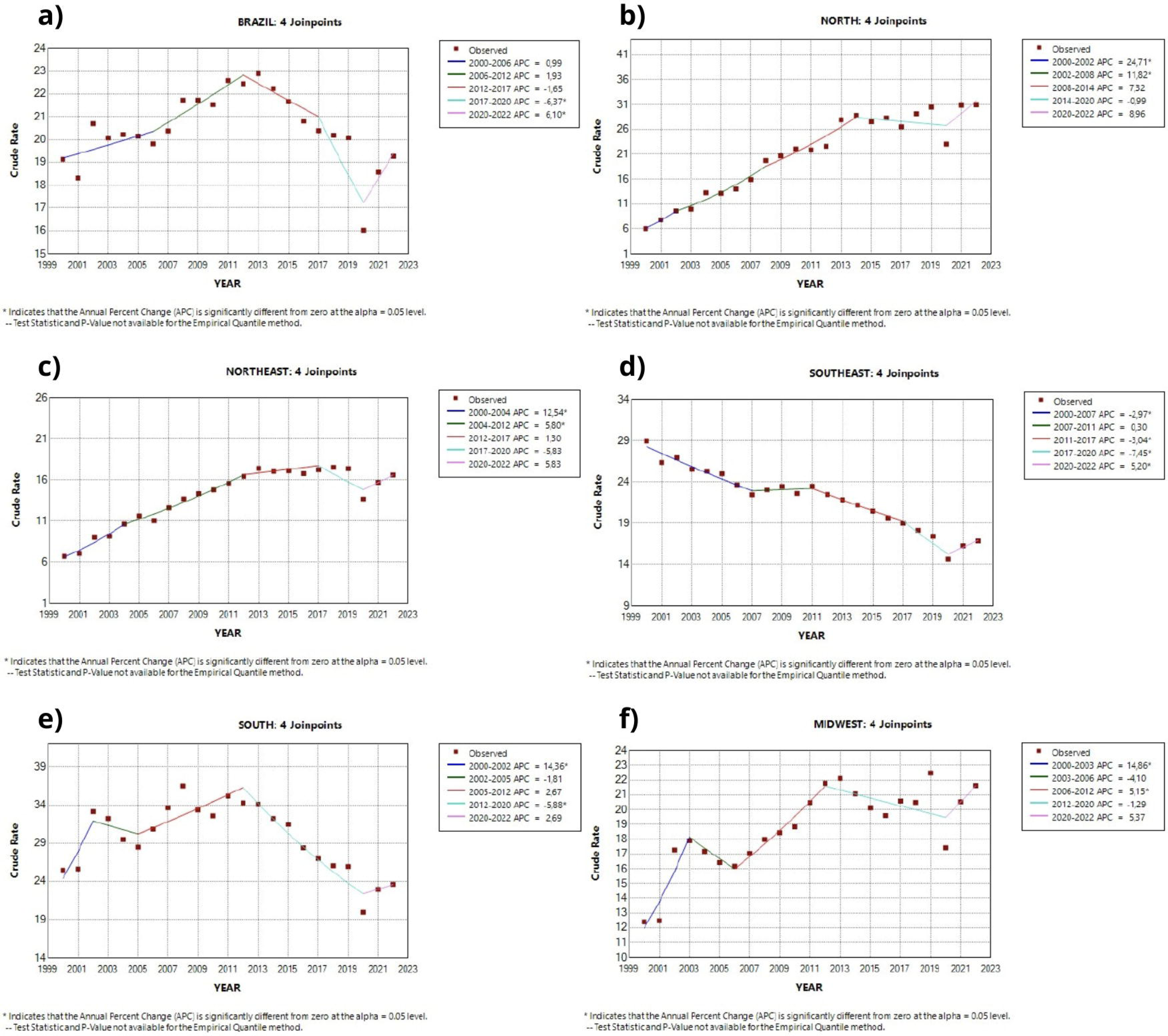
Trend analysis of HIV infection in Brazil and its regions, from 2000 to 2022. Brazil, 2025.

In contrast, the North (Fig. 2b) and Northeast (Fig. 2c) regions showed a gradual increase throughout the analyzed time series. The Southeast, which started in 2000 with the highest incidence rate of 29 cases per 100,000 inhabitants—approximately four times higher than the North and Northeast regions in the same period—presented one of the lowest rates in the country by 2022 (Fig. 2d).

**Fig. 2.**
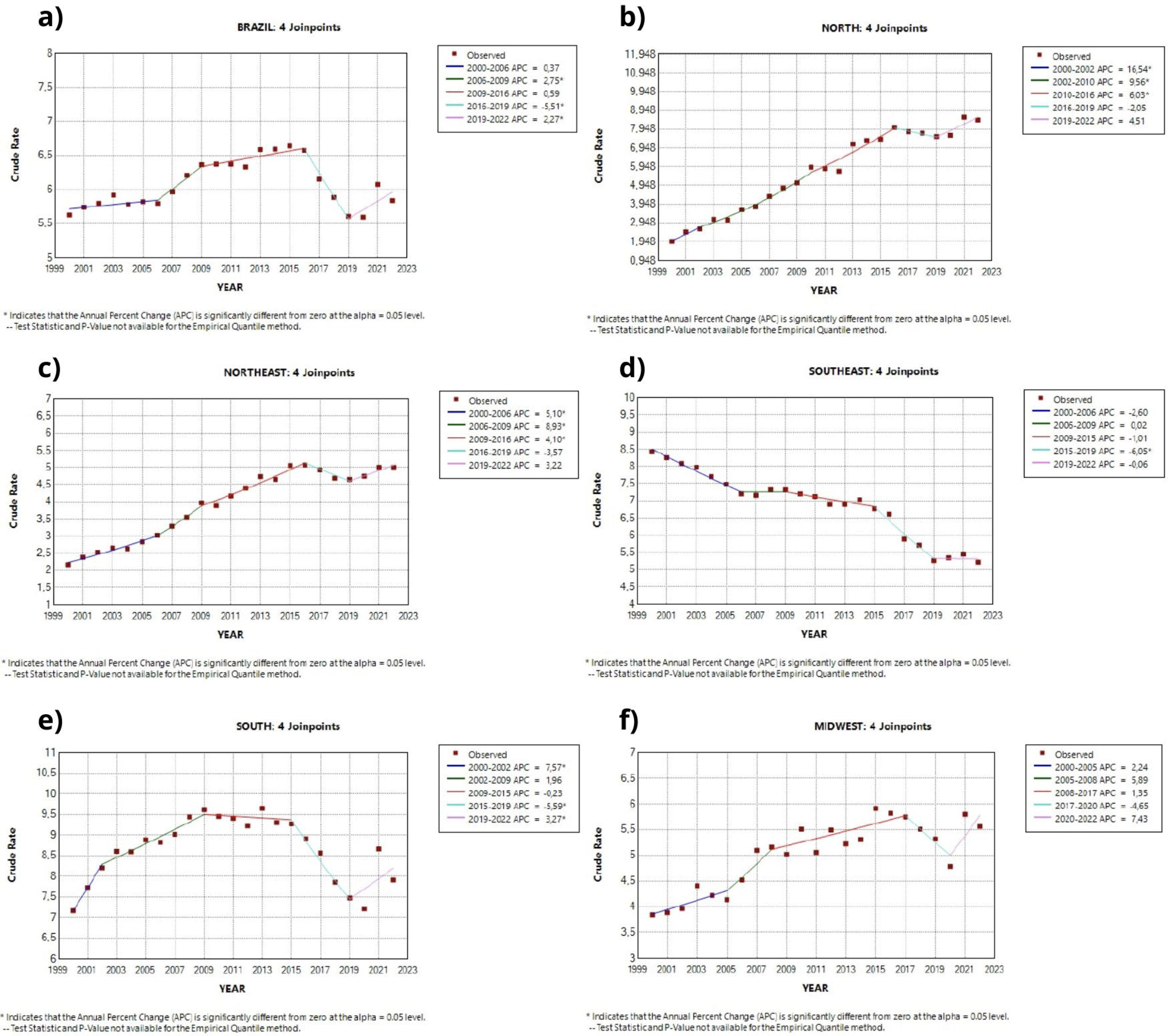
Trend analysis of HIV mortality in Brazil and its regions, from 2000 to 2022. Brazil, 2025.

Regarding the South, this region exhibited a pattern most similar to the national scenario, with an increasing curve until 2012, followed by a gradual and significant reduction (APC = −5.88), with variation during the pandemic period (Fig. 2e). The Central-West region showed growth until 2012, followed by a decreasing trend (APC = −1.29), albeit more discreetly, and an inversion of the pattern at the end of the period (APC = 5.37), indicating a tendency toward increase (Fig. 2f).

Table 1 presents the detailed Joinpoint® regression analysis for HIV incidence rates in Brazil and its regions. It is possible to observe that all regions demonstrated significant APCs in at least two line segments, i.e., at least two distinct periods within the analyzed time series. Notably, there was a marked increase in incidence across all regions between 2000 and 2002, except for the Southeast region, which showed a significant and decreasing trend during nearly the entire period.

**Table 1.**
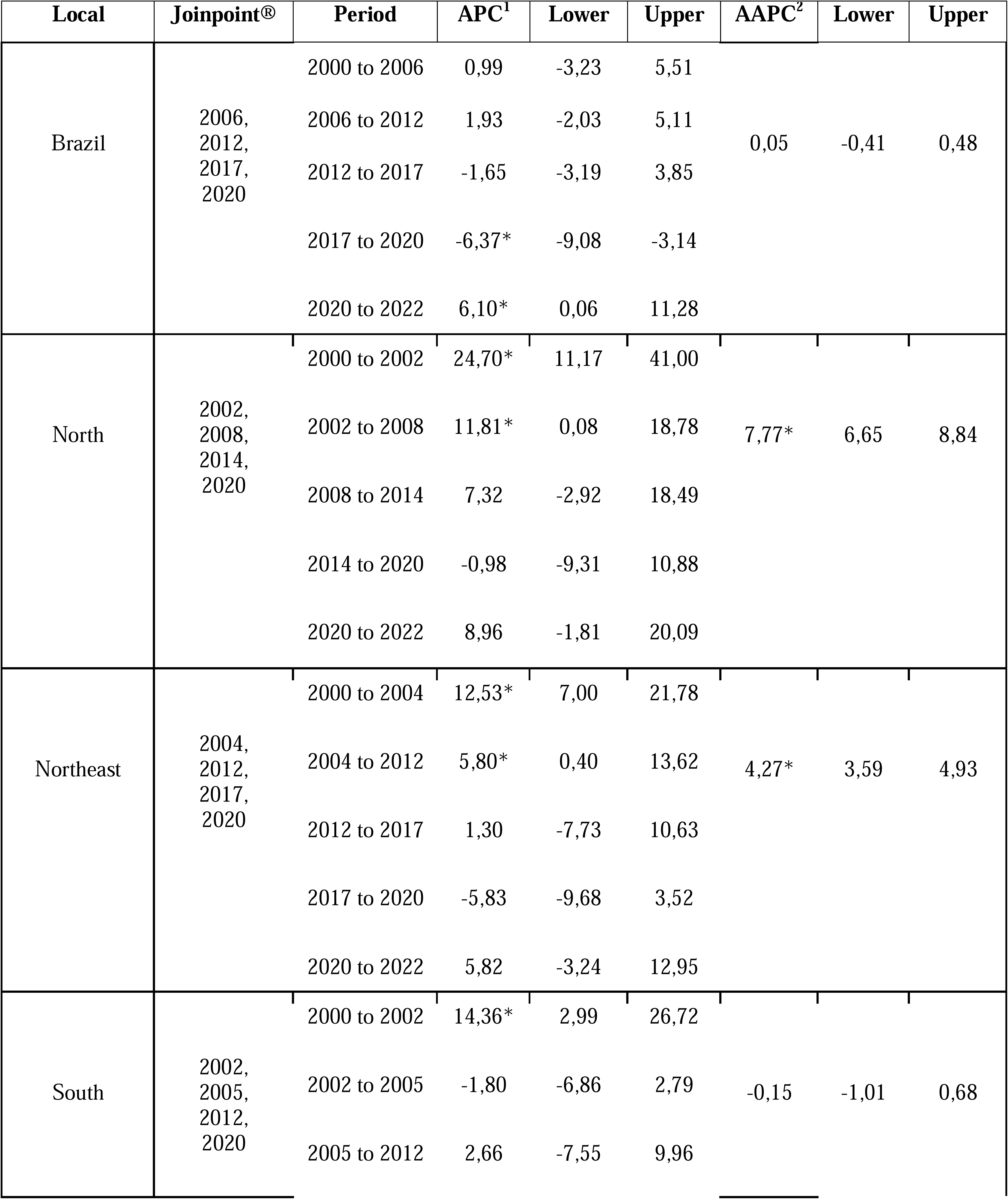
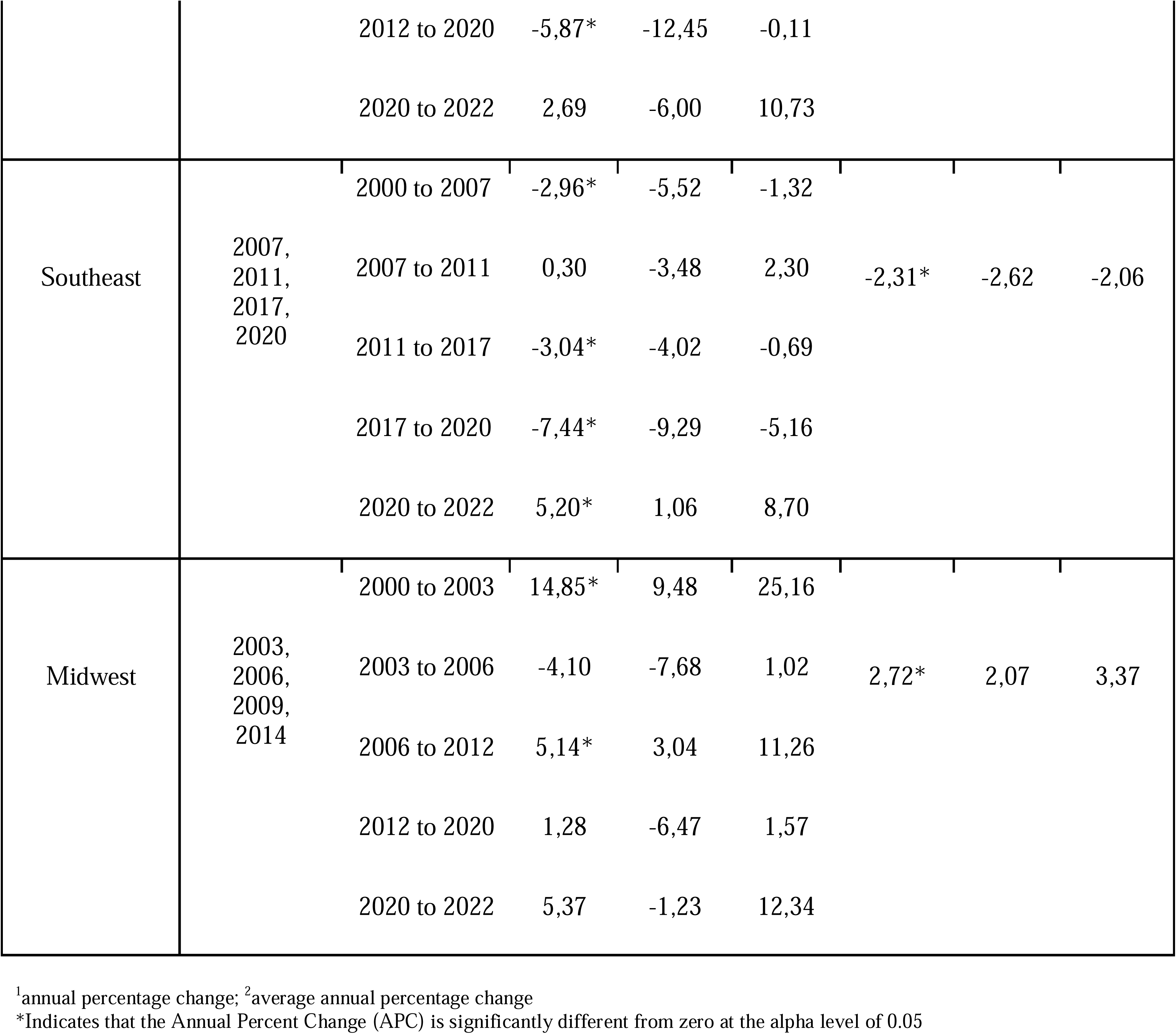
Joinpoint® analysis of HIV incidence in Brazil and its regions, 2011 to 2021. Brazil, 2025.

Figure 2a shows the HIV mortality rate in Brazil, highlighting an upward trend in the curve until 2016, reaching 6.6 deaths per 100,000 inhabitants. This is followed by a decrease until 2019 (APC = −5.51), and then a rise again during the pandemic (APC = 2.27). The North (Fig. 2b), Northeast (Fig. 2c), and Central-West (Fig. 2f) regions exhibit a consistently increasing trend, except for the period from 2016 to 2019, which showed a slight decrease (APC = −2.05, APC = −3.57, and APC = −4.65, respectively).

On the other hand, the Southeast region (Fig. 2d) is the only one exhibiting line segments with decreasing or stationary APCs throughout the analyzed period, indicating a positive trend in reducing HIV mortality, in contrast to the other Brazilian regions. The South region (Fig. 2e) showed a gradual increase until 2009, after which it experienced a stationary APC, followed by a significant reduction (APC = −5.59), but mortality rose again during the COVID-19 pandemic (APC = 7.43).

Table 2 presents a detailed Joinpoint® analysis of HIV mortality rates across Brazil. The growth pattern follows that of incidence, albeit more discreetly. Additionally, this increase is more pronounced during the first decade of the 21st century. Similar to incidence rates, the Southeast region deviates from the other regions’ behavior, demonstrating a reduction in mortality rates throughout most of the analyzed time series.

**Table 2.**
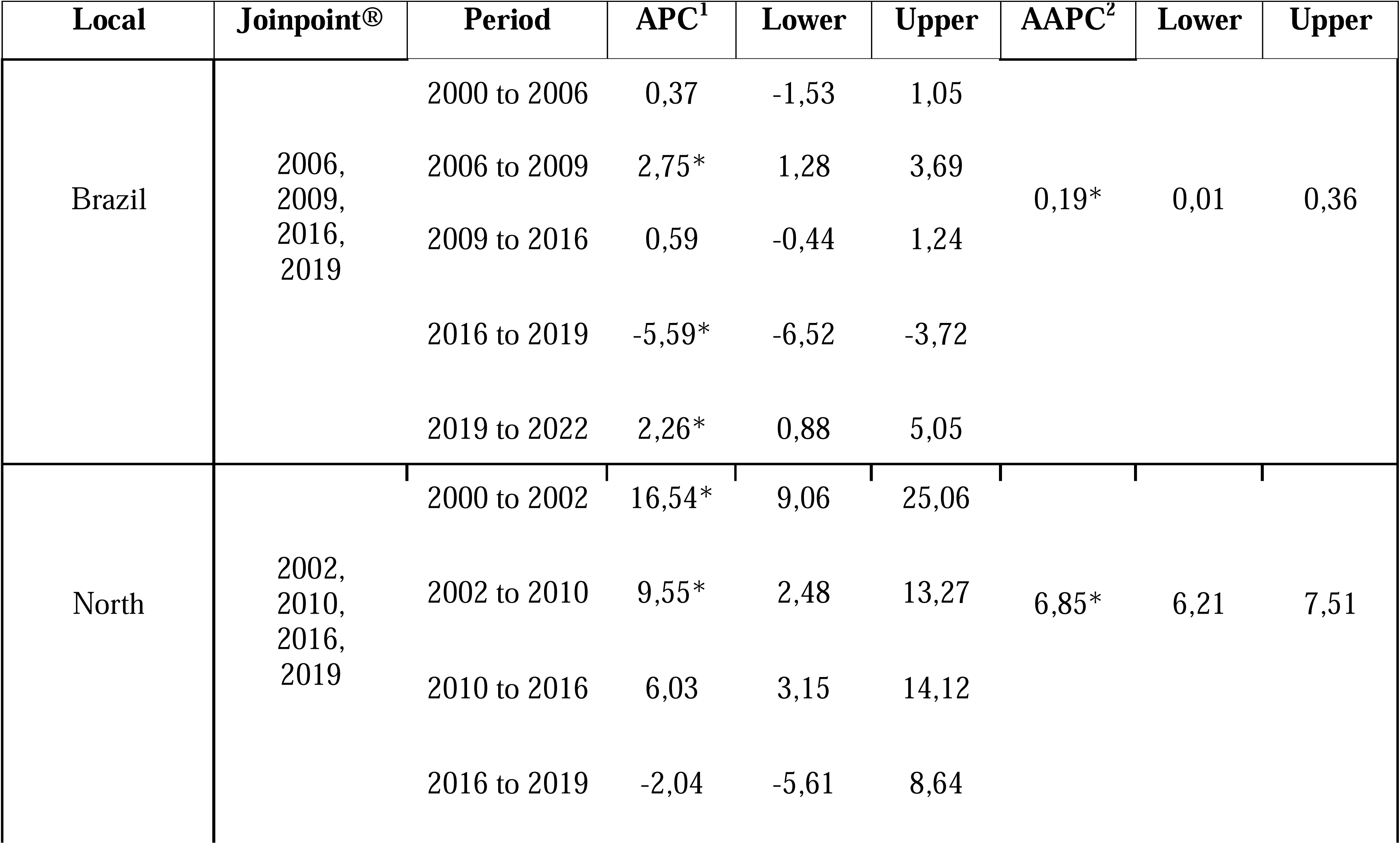
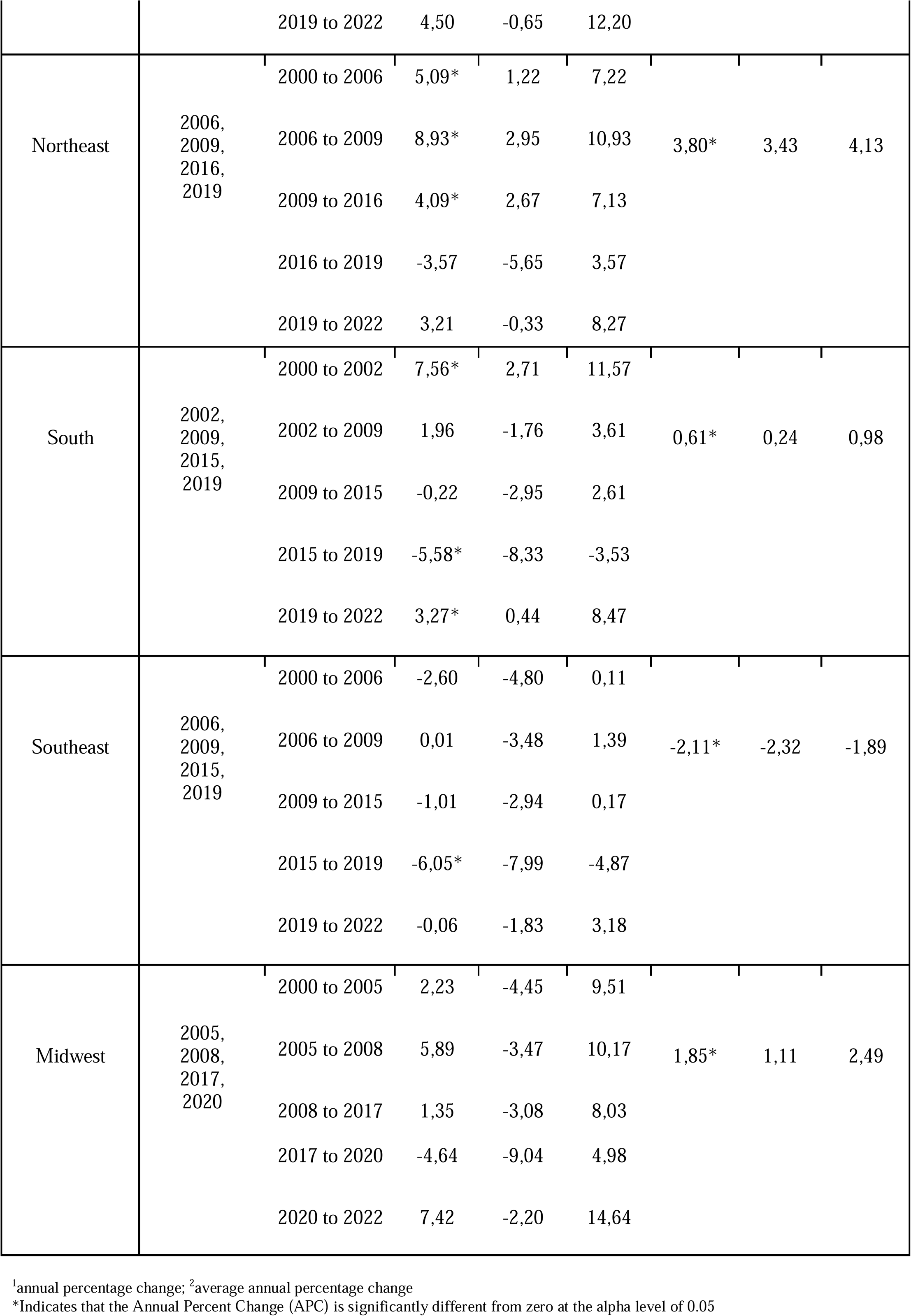
Joinpoint® analysis of HIV mortality in Brazil and its regions, 2011 to 2021. Brazil, 2025.

## Discussion

The temporal analysis revealed distinct patterns of HIV incidence and mortality across the Brazilian regions. While the country as a whole showed fluctuations over the study period, the North and Northeast regions exhibited a continued upward trend, whereas the Southeast demonstrated a consistent decline. The South and Central-West showed intermediate patterns. Mortality trends followed a similar trajectory to incidence, albeit less pronounced, with a notable sustained decrease in the Southeast. These findings highlight regional disparities and underscore the need for region-specific response strategies.

In certain disease types, health determinants and conditioning factors are especially important, considering the significant impact that living conditions have on the population’s illness process [16]. HIV falls within this context, being a virus strongly influenced by behavioral factors, access to health services, and recurrent violations of human rights, as is characteristic of discrimination against drug users, homosexuals, and sexual violence against women [17].

Brazil has provided antiretroviral treatment (ART) since 1996, challenging budget expenditures that were estimated at US$ 10,000 per patient, a significant amount for the public budget of a developing country [18]. Furthermore, in the same year, the National Congress promulgated Law No. 9,313/1996, guaranteeing free and universal access to medications for AIDS treatment [19]. The high cost to the health system is often seen as a barrier to reducing HIV indicators, given that it is a chronic disease requiring lifelong treatment.

Moreover, several economic strategies were fundamental in increasing the country’s competitiveness in purchasing these medications and reducing prices, including substantial investment in the production of national generics. These efforts culminated in a gradual increase in people living with HIV (PLHIV) using ART. Data indicate that in 1997 only 35,000 Brazilians were on ART, increasing to 185,000 by 2008 [18].

By 2010, nearly all PLHIV in wealthier countries had access to appropriate treatment, while only 25% had access in underdeveloped countries. In the first decade of the 21st century, it is estimated that 95% of incidence occurred in developing countries, yet access to medications remained scarce [20].

Meanwhile, Brazil stands out for promulgating its first policy to combat HIV, called the National Policy for STDs/AIDS, which aims to reduce the incidence of HIV/AIDS and other STDs; expand access to diagnosis, treatment, and care; and strengthen public and private institutions responsible for STD and AIDS control [21].

However, it is worth noting that treatment influences mortality reduction but does not impact the incidence. In the results of this study, a deceleration in mortality was observed from 2009 to 2016. Brazil had been experiencing growth with an APC = 2.75, which reduced to 0.59 during this period. Nonetheless, this deceleration was not uniform across all regions, as reductions in the North and Northeast were smaller.

This scenario may be related to difficulties in these regions regarding the lack of availability and access to health services, unfavorable socioeconomic determinants, and low quality of health services. Many variables are involved in the health-disease process, and socioeconomic inequalities can generate inequities in health [22,23].

Therefore, there is a fundamental need to invest in prevention methods such as male and female condoms, pre-exposure prophylaxis (PrEP), and post-exposure prophylaxis (PEP); harm reduction policies—such as needle exchange for injectable drug users—rapid testing; educational actions targeting diverse populations across various age groups and educational levels; and prioritization of pregnant women in prenatal care [24].

Another impactful factor is the high rate of rehospitalizations due to opportunistic infections associated with HIV. A study conducted in the Southern region of Brazil, which analyzed hospital readmission within 30 days, found that 11.5% of HIV-positive patients were readmitted less than 30 days after discharge [25].

Brazil’s efforts are part of the “Test and Treat” strategy; however, it is essential to consider that in a large, developing country like Brazil, with intrinsic regional disparities, it is important to also incorporate monitoring of prevention actions focused on structural and behavioral axes targeted at key populations [26].

Despite advances in combating HIV, many barriers remain regarding the distribution of health services and actions, which occur unevenly, especially in access to antiretrovirals, low quality of prenatal care, and difficulties in rapid testing [27].

In this context, the increase of HIV in the North and Northeast regions, as presented in the results of this study, may reflect behavioral changes caused by rapid urbanization without adequate efforts to reduce regional health inequities, difficulties in accessing services, chronic underfunding, stigma, and discrimination. A study conducted in Bahia, a state in the Northeast, reported a higher concentration of sexually transmitted infections in major cities of the state, suggesting that urbanization and urban agglomerations may be impacting this scenario [28].

Conversely, the sustained decline in HIV incidence and mortality in the Southeast may be associated with the robustness of the healthcare network, more consolidated public policies, and higher population education levels. The state of São Paulo, which has the largest population in the Southeast region and Brazil, stood out for pioneering the first state program to combat HIV/AIDS in 1993, even before the creation of the Unified Health System (SUS) [29].

Additionally, other factors such as broader and higher quality primary healthcare coverage, greater reach of specialized services (SAEs), early testing, and regular access to antiretroviral therapy (ART) make the Southeast stand out compared to other regions and have allowed it to reduce its indicators throughout the analyzed time series [30].

Furthermore, the abrupt nationwide reduction in notifications in 2020, the first year of the COVID-19 pandemic, reveals the vulnerability of surveillance systems in the face of health emergencies, directly impacting the continuity of care and early detection. This scenario of health system overload, with the urgent need to inject resources for a specific demand, highlights the risk of insufficient investment in HIV and other non-respiratory diseases in Brazil [31,32].

Although determinants and conditioning factors of health are important for epidemiology of community health conditions, one study indicates that State Health Plans across all Brazilian regions, developed between 2012 and 2015, do not clearly define what health determinants and conditioning factors are, nor do they present the theoretical model used in their analyses, which is essential for adopting criteria in constructing indicator-related analyses.

Despite improvements in health notification systems, there remains an evident disparity in health surveillance services across various locations in Brazil, demonstrated by variations in data coverage and validity [33].

Therefore, the need for grounding based on health conference deliberations stands out, as these are well aligned with social demands. It is also emphasized that the theoretical model used must be capable of reflecting the analyzed scenario, as well as the technical, political, and institutional capacity permeating the health system.

This study presents some limitations inherent to ecological studies based on secondary data, particularly related to underreporting and data entry errors. Furthermore, the use of macro-regional analysis may hinder the identification of important nuances between states and micro-regions, and it does not allow for the detection of categorical changes within the studied population. Therefore, further studies are recommended to explore the geographic characteristics and the profile of PLHIV in greater detail.

## Conclusion

The study analyzed temporal trends in HIV incidence and mortality in Brazil and its regions. Nationally, HIV incidence and mortality exhibit an overall declining pattern. However, this trend is not uniformly observed across all regions of the country, suggesting that demographic and socioeconomic determinants may be playing a significant role, particularly in sustaining structural health inequities.

The findings highlighted that the Southeast region has been more effective in curbing the spread and mortality of HIV, with the state of São Paulo serving as a pioneer in the development of targeted health policies for this condition. In contrast, the North and Northeast regions have shown a gradual and consistent increase in HIV incidence and mortality rates, indicating a divergence from the national trend.

This scenario underscores the need for public policies focused on key and priority populations, while also taking into account the specific profiles and needs of each region. Furthermore, it is essential to recognize AIDS as a complex syndrome that requires comprehensive strategies involving harm reduction, prophylaxis, and treatment.

## Data Availability

If the data are held or will be held in a public repository, include URLs, accession numbers or DOIs. If this information will only be available after acceptance, indicate this by ticking the box below. For example: All files are available from the DATASUS database (link: https://datasus.saude.gov.br/informacoes-de-saude-tabnet/).

## Notes

### Competing Interest Statement

The authors have declared no competing interest.

### Funding Statement

The author(s) received no specific funding for this work.

